# Clonal Hematopoiesis of Indeterminate Potential Refines Cardiovascular Risk Stratification in Cardiovascular-Kidney-Metabolic Syndrome Stages 0–3

**DOI:** 10.64898/2026.06.04.26354963

**Authors:** Jian Lu, Shuaigang Sun, Zekai Deng, Shunwei Wang, Chenping Wei, Shimin Jiang, Wenge Li

## Abstract

**Background:** Chronic low-grade inflammation drives cardiovascular-kidney-metabolic (CKM) syndrome. Clonal hematopoiesis of indeterminate potential (CHIP), an age-related driver of systemic inflammation, is linked to several cardiometabolic disorders. However, whether CHIP modifies CKM progression and contributes to heterogeneity in cardiovascular disease (CVD) risk within the CKM framework remains uninvestigated.

**Methods:** This cohort study included 307,025 UK Biobank participants at CKM stages 0–3 free of baseline CVD. CHIP status was identified via whole-exome sequencing (WES). The association between CHIP and baseline CKM severity was examined, along with the independent and joint effects of CHIP and CKM stages on incident CVD risk. The joint effects of CHIP and polygenic risk scores (PRS) were further assessed, and the incremental predictive value of incorporating CHIP into the AHA PREVENT equations was evaluated.

**Results:** CHIP carriers were more likely to present with advanced CKM stages [OR 1.14 (1.09-1.20), *P* < 0.001] and exhibited higher incident CVD risk during follow-up [HR 1.13 (1.08-1.18), *P* < 0.001]. Significant joint effects between CHIP and CKM stages were observed, with the highest risk among CHIP carriers at CKM stage 3 [HR 1.63 (1.50-1.78), *P* < 0.001]. Large or multiple CHIP mutations conferred greater hazards, with distinct gene-specific effects observed. Moreover, CHIP and high genetic risk also jointly amplified CVD susceptibility. Most importantly, incorporating CHIP into AHA PREVENT significantly improved risk discrimination.

**Conclusions:** CHIP is a significant risk factor associated with more advanced CKM stages and amplifies incident CVD risk. Integrating CHIP into existing prevention strategies may refine CVD risk stratification.

**Graphic Abstract:** 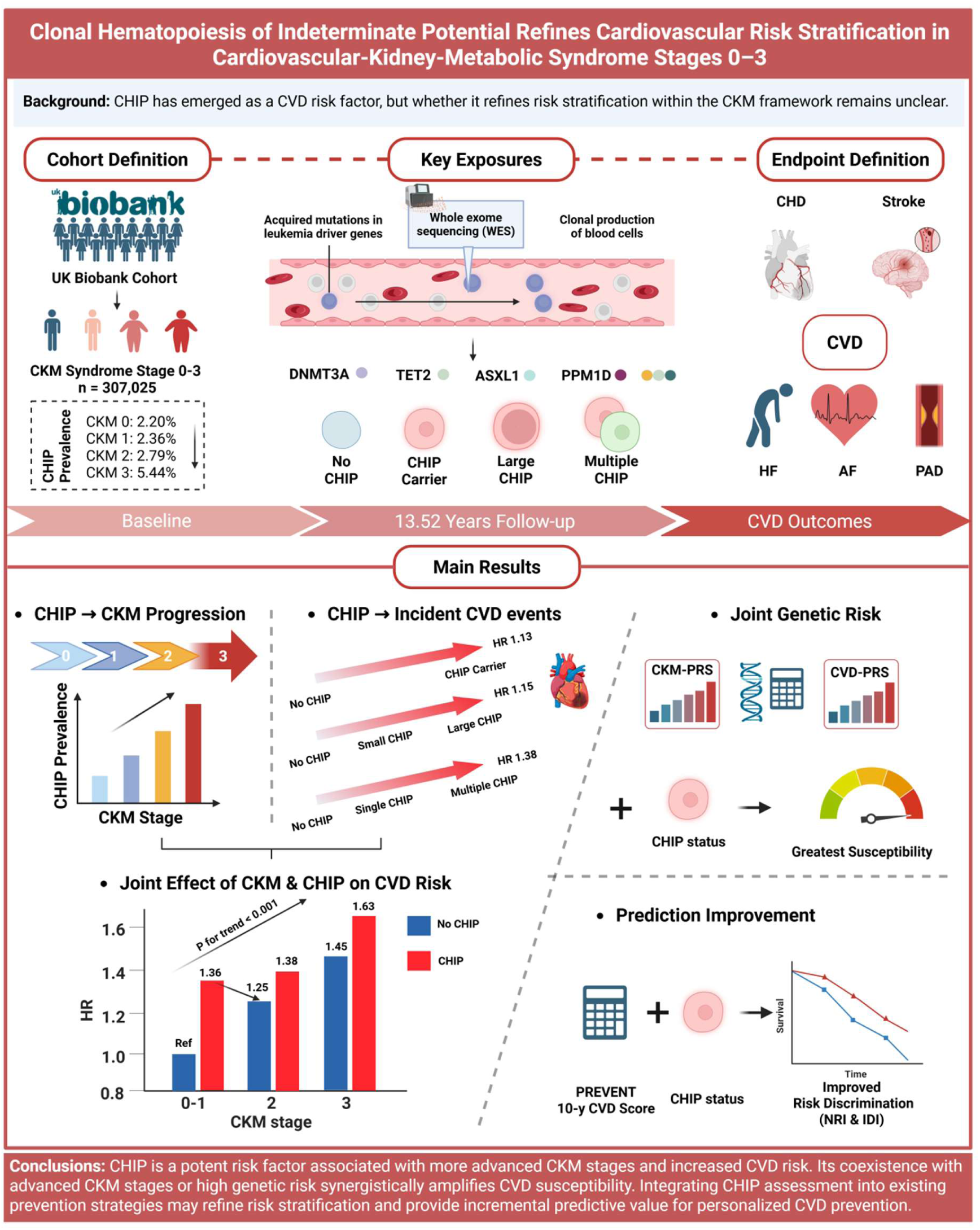

**Clinical Perspective:** *What Is New?:*

- This study provides large-scale evidence that clonal hematopoiesis of indeterminate potential (CHIP) is associated with more advanced cardiovascular-kidney-metabolic (CKM) stages and jointly amplifies future cardiovascular disease (CVD) risk within the CKM staging framework.
- Somatic CHIP and germline polygenic susceptibility were jointly associated with CKM severity and CVD risk, with the greatest risk observed among individuals with both high polygenic risk and CHIP.

*What Are the Clinical Implications?:*

- CHIP significantly refines CVD risk stratification across AHA PREVENT risk categories and provides incremental predictive value. Incorporating CHIP into CKM-based prevention strategies may help identify individuals with excess residual cardiovascular risk before the development of clinical CVD.

## 1. Introduction

Cardiovascular disease (CVD) remains the leading cause of mortality worldwide ^[1]^, with its escalating burden largely driven by the widespread prevalence of modifiable risk factors such as obesity, diabetes, and chronic kidney disease (CKD) ^[2]^. In 2023, the American Heart Association (AHA) formally introduced the concept of cardiovascular-kidney-metabolic (CKM) syndrome, a multisystem framework that stratifies individuals from stage 0 (no risk factors) to stage 4 (established clinical CVD), emphasizing the progressive nature of the condition and the synergistic interplay among metabolic dysfunction, renal impairment, and cardiovascular abnormalities ^[3]^. It aims to shift the clinical focus toward integrated and preventive disease management ^[4]^. Notably, CKM stages 0–3 represent a critical preclinical window, encompassing over 90% of the general adult population ^[5]^. Although the CKM staging system provides a robust blueprint for stratified metabolic management, significant heterogeneity in CVD risk persists among individuals within the same stage, necessitating the identification of novel biological drivers to further refine risk stratification.

Clonal hematopoiesis of indeterminate potential (CHIP) is characterized by the clonal expansion of hematopoietic cells harboring somatic mutations in a restricted set of leukemia-associated driver genes (e.g., *DNMT3A, TET2, ASXL1, and JAK2*) in the absence of hematological malignancy ^[6]^. CHIP is a common age-related condition, with a prevalence that increases substantially with age, affecting at least 10–20% of individuals over 70 years ^[7]^. Furthermore, CHIP has been identified as a potent and independent risk factor for various CVDs, such as coronary heart disease (CHD) ^[8,9]^, heart failure (HF) ^[10]^ and arrhythmias ^[11]^, primarily driven by the hyperactivation of NLRP3 inflammasome and the enhanced synthesis of pro-inflammatory cytokines such as interleukin (IL)-1β and IL-6, which accelerate atherosclerotic plaque formation and promote myocardial inflammation and fibrosis ^[12]^. Given that both aging and inflammatory processes are central to CHIP and CVD risk, we hypothesize that CHIP may act as a biological amplifier within the CKM framework. Such amplification could not only accelerate the transition from early-stage metabolic dysfunction to advanced CKM stages but also partially explain the residual CVD risk heterogeneity observed among individuals within the same CKM stratum.

Therefore, leveraging whole-exome sequencing (WES) data from a large-scale UK Biobank cohort, this study aims to systematically investigate whether and how CHIP interacts with CKM stages to jointly influence the risk of incident CVD and its components, and to determine whether incorporating CHIP significantly improves the performance of existing CVD prediction methods. Furthermore, by integrating genome-wide association studies (GWAS) and polygenic risk scores (PRS), we aim to provide an integrated evidence base to inform more personalized prevention strategies.

## 2. Methods

### 2.1 Study population and data source

This study used data from the UK Biobank, a prospective cohort of over 500,000 participants aged 37 to 73 years recruited between 2006 and 2010 from 22 assessment centers across England, Scotland, and Wales. Participants completed a questionnaire, underwent physical examination, and provided blood samples for WES or whole-genome sequencing (WGS). Detailed descriptions of the data collection procedures are available elsewhere ^[13]^. The study was approved by the North West Multi-centre Research Ethics Committee (MREC) (21/NW/0157), with all participants providing written informed consent.

Among 450,679 participants with available WES data, we excluded those lacking data required for CKM definition (n = 98,797), those with missing covariates (n = 12,311), and those with baseline CVD (n = 32,546), resulting in 307,025 participants at CKM stages 0–3 for analysis (**Figure S1**). Details regarding missing information for CKM-defining variables are provided in **Table S1**. GWAS summary data for CVD outcomes were obtained from the FinnGen consortium (**Table S2**).

### 2.2 Definition of CKM syndrome

The components of CKM syndrome included excess or dysfunctional adiposity, metabolic risk factors, chronic kidney disease (CKD), and CVD. Stages were defined at baseline according to the AHA Presidential Advisory framework ^[4]^ and adapted for the UK Biobank (**Table S3**): Stage 0 (no risk factors), Stage 1 (excess/dysfunctional adiposity), Stage 2 (metabolic risk factors or moderate- to high-risk CKD), Stage 3 (very high-risk CKD or 10-year CVD risk ≥10%), and Stage 4 (established clinical CVD). Participants with Stage 4 at baseline were excluded. Estimated glomerular filtration rate (eGFR) was calculated using the 2021 CKD-EPI creatinine-cystatin C equation ^[14]^. Detailed criteria for the AHA Predicting Risk of CVD EVENTs (PREVENT) equations ^[15]^ and KDIGO-based CKD risk stratification ^[16]^ are provided in **Table S4 and S5**.

### 2.3 Assessment of CHIP

CHIP was identified via whole blood-derived WES data (Illumina NovaSeq 6000, Regeneron Genetics Center) ^[17,18]^. Somatic variants were detected using Mutect2 within the Genome Analysis Toolkit ^[19]^ and curated as previously described ^[17]^ using a refined list of 58 genes (**Table S6**). Variants with total read depth <20, alternate allele depth <5, or insufficient strand support were excluded ^[17]^.

CHIP was defined as the presence of at least one mutation with variant allele fraction (VAF) ≥2%. Exposures included: (1) CHIP carrier status; (2) CHIP VAF (small: 2% ≤ VAF < 10%; large: VAF ≥ 10%); (3) CHIP mutations (single and multiple, ≥ 2). For multiple mutations, the highest VAF was used. Gene-specific analyses focused on common driver genes (*DNMT3A, TET2, ASXL1,* and *JAK2*), DNA damage repair genes (*PPM1D* and *TP53*), and spliceosome genes (*PRPF8, SF3B1, SRSF2, U2AF1,* and *ZRSR2*). The Clonal Hematopoiesis Risk Score (CHRS) was calculated per established methodology (**Table S7**) ^[20]^ and categorized as low (< 9.5), intermediate (10.0–12.0), or high (≥ 12.5).

### 2.4 Assessment of covariates

Baseline covariates included age, sex, ethnicity, educational attainment, alcohol consumption, smoking status, sleep duration, assessment center and Townsend Deprivation Index (TDI). Body Mass Index (BMI) was calculated from measured height and weight. Physical activity was considered adequate if participants engaged in ≥ 150 minutes of moderate-intensity or ≥ 75 minutes of vigorous-intensity activity per week, or an equivalent combination ^[21]^. Diet quality was assessed using a standardized scoring system based on the intake of seven dietary components (**Table S8**) ^[22]^. Furthermore, medical history and medication use (antihypertensive, antidiabetic, lipid-lowering) were obtained from nurse-led interviews (**Table S9**).

### 2.5 Assessment of CVD

The primary outcome was the incidence of a composite CVD endpoint, including coronary heart disease (CHD), stroke, heart failure (HF), atrial fibrillation (AF), and peripheral artery disease (PAD), consistent with the AHA criteria for CKM Stage 4. Participants at CKM stage 0-3 free of CVD at baseline were followed from the date of recruitment until the first incident CVD event, death, loss to follow-up, or the end of the follow-up period (November 30, 2022), whichever occurred first. Incident cases were identified through linkage with hospital inpatient records, primary care data, and national death registries. Detailed mapping of the International Classification of Diseases, Tenth Revision (ICD-10) codes, self-reported data, and “first occurrence” fields used to define each outcome are provided in **Table S10**.

### 2.6 Genome-wide association study for CKM syndrome

Genotyping was performed using the Applied Biosystems UK BiLEVE Axiom Array by Affymetrix and the Applied Biosystems UKB Axiom Array. In this study, we utilized version 2 for directly genotyped variants and version 3 for imputed genotypes. Quality control (QC) procedures were applied to the directly genotyped autosomal variants using PLINK2, and variants with a minor allele frequency (MAF) < 1%, genotyping call rate < 90%, or Hardy-Weinberg equilibrium *P* < 1 × 10^-15^ were excluded. Further linkage disequilibrium (LD) pruning was performed using a window size of 1,000 variants, a step size of 100, and an r^2^ threshold of 0.1.

For the final analysis, the study population was randomly split into equal training (50%) and testing (50%) sets. The GWAS was conducted in the training set (n = 153,512) using REGENIE (v3.2.1), a two-step whole-genome regression method that accounts for sample relatedness and population structure ^[23]^. In REGENIE Step 1, the genetic relationship matrix was estimated using the QC-filtered directly genotyped data. In REGENIE Step 2, association testing was performed on the imputed dataset across chromosomes 1–22, restricting the analysis to variants with high imputation confidence (INFO score ≥ 0.8) and a minor allele count (MAC) ≥ 10. The stages of CKM syndrome were modeled as an ordinal quantitative trait (values 0, 1, 2, 3). All models were adjusted for age, sex, genotyping array, and the top ten principal components. Quantile-Quantile (Q-Q) plots and the genomic inflation factor (λGC) were utilized to assess potential population stratification or technical bias.

### 2.7 Construction of polygenic risk score

For CKM syndrome, we utilized the GWAS summary statistics derived from our internal training set (n = 153,512), while for CVD components, SNP weights were calculated using external publicly available GWAS summary data (**Table S2**). The posterior SNP effect sizes were estimated using PRS-CS, a Bayesian approach that employs a continuous shrinkage prior to account for LD patterns based on the 1000 Genomes Project European reference panel ^[24]^. A global shrinkage parameter of φ= 10^-2^ was applied across all autosomes. These weights were subsequently applied to the testing set (n = 153,513) to generate individual-level standardized PRS.

### 2.8 Statistical Analysis

Baseline characteristics were presented as mean (SD), median [IQR], or n (%). Ordinal logistic regression and Cox proportional hazards models estimated odds ratios (ORs) and hazard ratios (HRs) with 95% confidence intervals (CIs) for associations between CHIP status, CKM stages, and incident CVD. Models were stepwise adjusted for age, sex, ethnicity, BMI, education, TDI, smoking status, alcohol consumption, sleep duration, physical activity, diet quality, assessment center, hypertension, diabetes, and use of antihypertensive, antidiabetic, and lipid-lowering medications. The proportional hazards assumption was verified using Schoenfeld residuals, and the cumulative incidence of CVD was visualized using Kaplan–Meier curves. Sensitivity analyses included: (1) excluding CVD events within the first 2 or 5 years; (2) excluding deaths within 2 years; (3) excluding participants with urinary albumin below the detection limit (< 6.7 mg/L); and (4) additional adjustment for the PREVENT 10-year CVD risk score. Subgroup analyses were utilized to evaluate potential effect modification by various factors such as sex, age, and CKM stages. Joint exposure groups were constructed to evaluate combined effects of CHIP and CKM stages, as well as CHIP and genetic risk (based on median PRS). Additionally, the incremental predictive value of incorporating CHIP into PREVENT was quantified using the Net Reclassification Improvement (NRI) and the Integrated Discrimination Improvement (IDI). All analyses used R version 4.3.2, with two-sided *P* < 0.05 considered significant.

## 3. Results

### 3.1 Baseline characteristics

A total of 307,025 participants with CKM syndrome stage 0-3 were included (**Table 1**). The mean age at baseline was 56.00 (8.07) years, and 44.69% were male. Over a median follow-up of 13.52 (12.65-14.27) years, a total of 41,152 participants (13.40%) developed incident CVD events. Notably, participants with more advanced CKM stages exhibited significantly elevated rates of CHIP carrier, large CHIP, and multiple CHIP mutations (*P* < 0.001). Furthermore, those were also associated with increased rates of incident CVD events (*P* < 0.001). Other baseline characteristics, including age, sex, ethnicity, TDI, educational attainment, smoking status, alcohol consumption, sleep duration, physical activity, and all CKM components, varied across the four CKM stages. Additionally, **Table S11** presents baseline and prognostic characteristics stratified by the absence or presence of CHIP. Consistently, CHIP carriers were associated with a markedly higher risk of incident CVD events (*P* < 0.001).

**Table 1.**
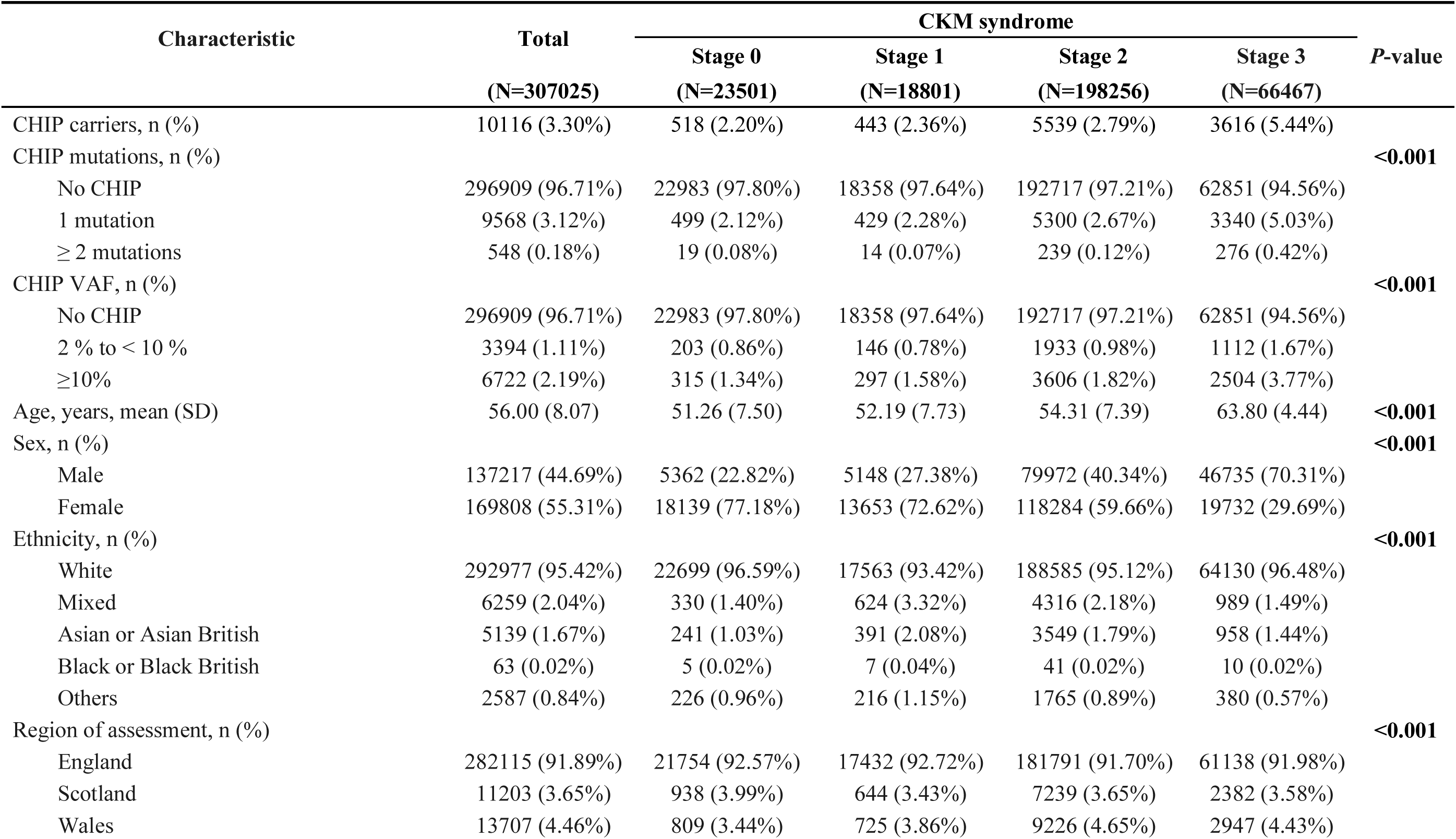
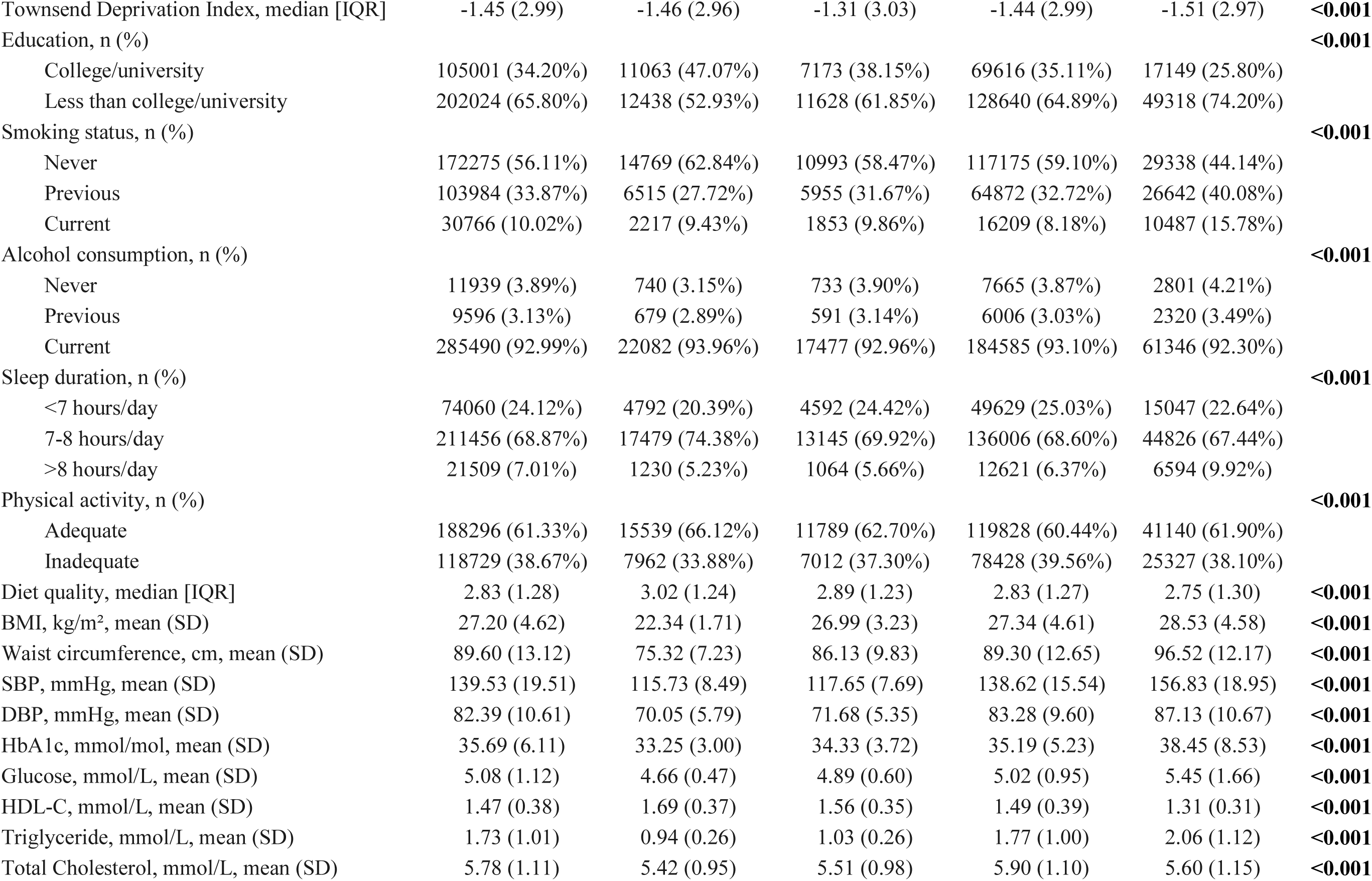
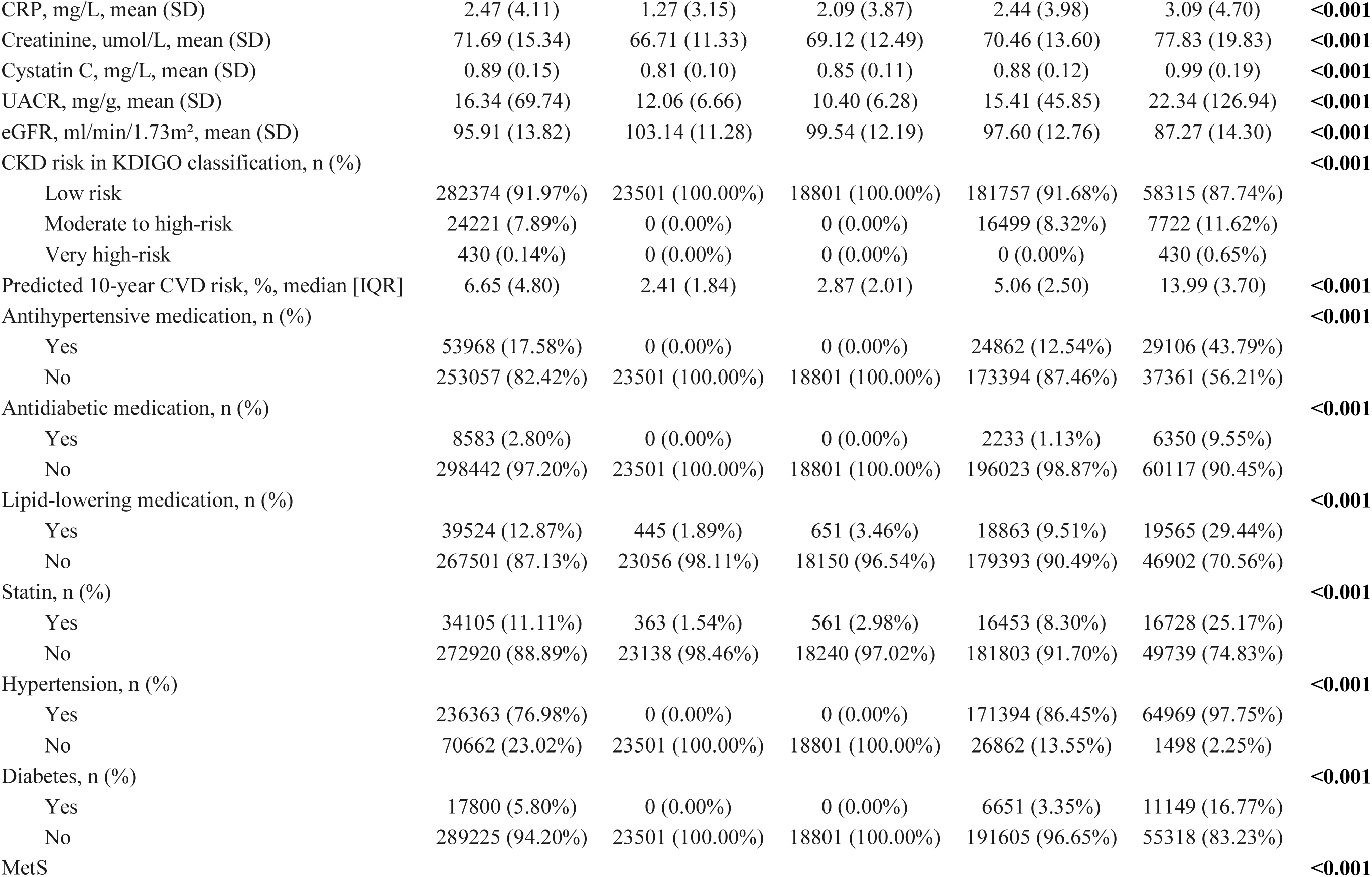
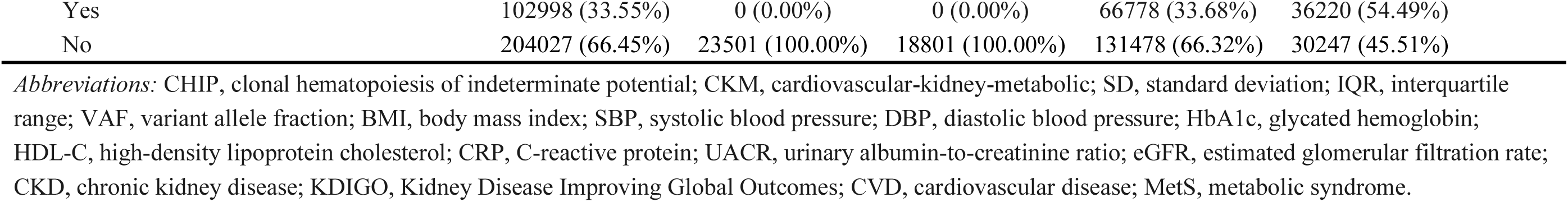
Baseline characteristics of participants according to CKM stage.

### 3.2 CHIP status and CKM syndrome

The prevalence and clonal burden of CHIP increased significantly with advancing CKM syndrome stages at baseline. As illustrated in **Figure 1A**, among the total population, the proportion of CHIP carriers rose from 2.2% in stage 0 to 5.4% in stage 3. Similarly, the prevalence of large CHIP increased from 1.3% to 3.8%, and multiple CHIP mutations rose from 0.1% to 0.4% (all *P* for trend < 0.001). When restricted to CHIP carriers, VAF values increased progressively as CKM stages advanced (Kruskal-Wallis *P* < 0.001). The proportion of large CHIP among carriers increased from 60.8% in stage 0 to 69.2% in stage 3, and the frequency of multiple mutations rose from 3.7% to 7.6% (all *P* for trend < 0.001). Age-stratified analyses further demonstrated that the prevalence of any CHIP, large CHIP, and multiple CHIP mutations rose steadily with biological age (**Figure 1B**), a trend that was consistently observed both in the total population and within each CKM stage (**Figure S2**).

**Figure 1.**
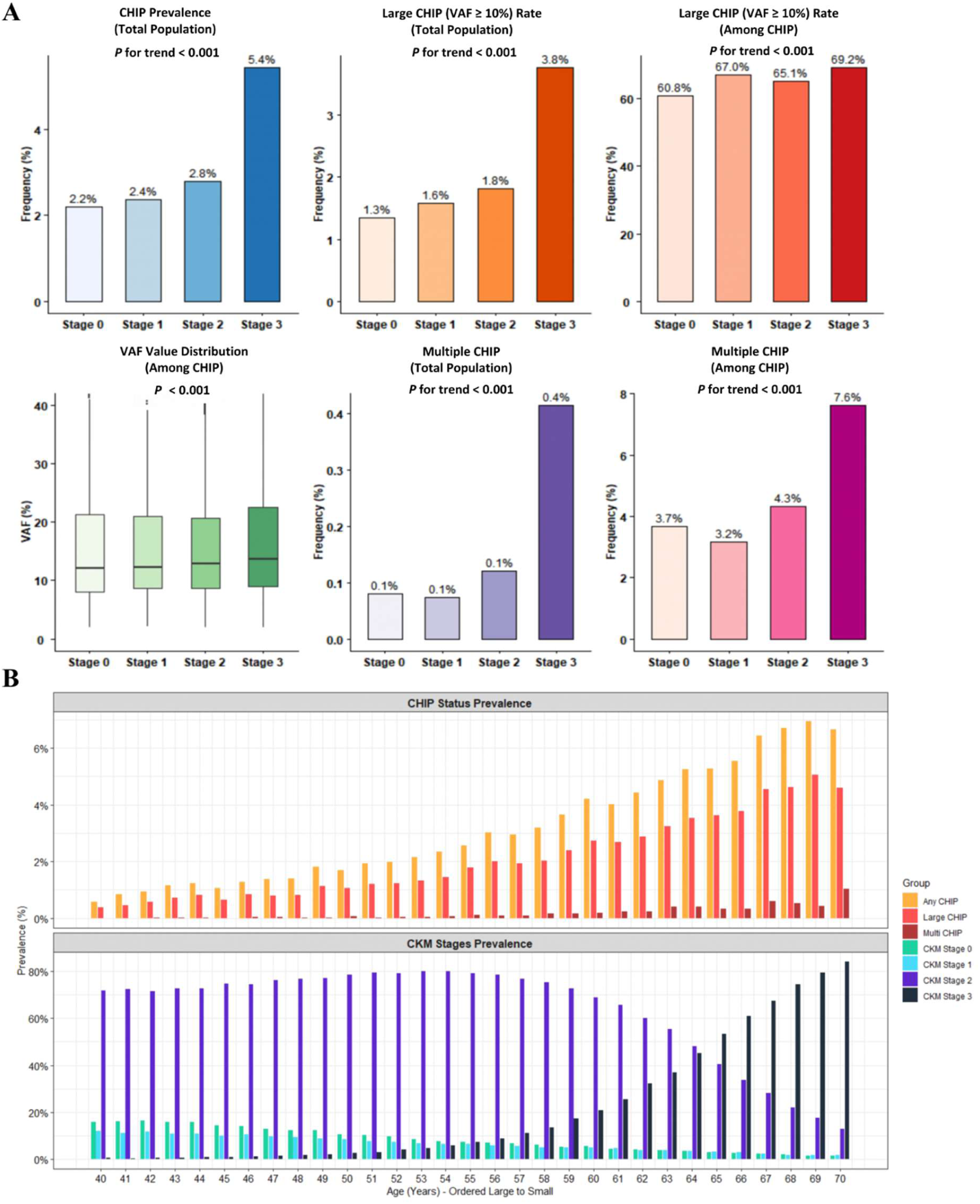
The prevalence and characteristics of CHIP status at baseline. (A) Prevalence of CHIP status across CKM stages 0 to 3. (B) Age-stratified distribution of CHIP status and CKM syndrome stages. CHIP, clonal hematopoiesis of indeterminate potential; CKM, cardiovascular-kidney-metabolic; VAF, variant allele fraction.

After full adjustment for covariates, ordinal logistic regression analyses revealed that CHIP carrier status was significantly associated with a higher risk of harboring a more advanced CKM stage at baseline compared to non-carriers [OR 1.14 (1.09-1.20), *P* < 0.001] (**Table 2**). Furthermore, significant dose-response relationships were also observed across different CHIP statuses. Specifically, participants with large CHIP [OR 1.18 (1.11-1.26), *P* < 0.001] and those with multiple mutations [OR = 1.48 (1.20-1.81), *P* < 0.001] exhibited a substantially increased risk of more severe CKM stages compared to those with no CHIP, small CHIP, or single CHIP, respectively (all *P* for trend < 0.001).

**Table 2.** Association of CHIP status with the risk of progression to advanced CKM syndrome stages.

| CHIP status | Model 1 |  | Model 2 |  | Model 3 |  |
| --- | --- | --- | --- | --- | --- | --- |
|  | OR (95% CI) | P value | OR (95% CI) | P value | OR (95% CI) | P value |
| CHIP carrier | 1.17 (1.12-1.22) | **<0.001** | 1.15 (1.10-1.20) | **<0.001** | 1.14 (1.09-1.20) | **<0.001** |
| CHIP VAF |  |  |  |  |  |  |
| No CHIP | Ref |  | Ref |  | Ref |  |
| Small CHIP (2% < VAF < 10%) | 1.07 (0.99-1.15) | 0.071 | 1.06 (0.98-1.14) | 0.158 | 1.06 (0.98-1.16) | 0.154 |
| Large CHIP (VAF ≥ 10%) | 1.22 (1.16-1.29) | **<0.001** | 1.20 (1.13-1.27) | **<0.001** | 1.18 (1.11-1.26) | **<0.001** |
| P for trend |  | **<0.001** |  |  |  | **<0.001** |
| CHIP mutations |  |  |  |  |  |  |
| No CHIP | Ref |  | Ref |  | Ref |  |
| Single CHIP | 1.15 (1.10-1.20) | **<0.001** | 1.13 (1.08-1.18) | **<0.001** | 1.12 (1.07-1.18) | **<0.001** |
| Multiple CHIP | 1.57 (1.30-1.88) | **<0.001** | 1.53 (1.26-1.85) | **<0.001** | 1.48 (1.20-1.81) | **<0.001** |
| P for trend |  | **<0.001** |  |  |  | **<0.001** |

*Abbreviations:* CHIP, clonal hematopoiesis of indeterminate potential; CVD, cardiovascular disease; OR, odds ratio; CI, confidence interval; VAF, variant allele fraction; Ref, reference group.
Model 1 was adjusted for age, sex, and ethnicity.
Model 2 was further adjusted for BMI, education, Townsend deprivation index, smoking status, alcohol consumption, sleep duration, physical activity, diet quality, and assessment center.
Model 3 was further adjusted for hypertension, diabetes, and use of antihypertensive, antidiabetic, and lipid-lowering medications.

### 3.3 CHIP status and CVD risk

As shown in **Figure 2**, during follow-up, the cumulative incidence of CVD and its components was significantly higher among CHIP carriers compared to non-carriers (log-rank *P* < 0.0001). Furthermore, the differences remained highly significant when participants were further stratified by CHIP VAF (large, small, or no CHIP) and CHIP mutations (multiple, single, or no CHIP) (all log-rank *P* < 0.0001). After full adjustment for covariates, Cox proportional hazards analyses revealed that CHIP carrier status was associated with a 13% increased risk of incident CVD compared to non-carriers [HR = 1.13 (1.08-1.18), *P* < 0.001]. Besides, large CHIP and multiple CHIP were associated with a 15% and 38% higher risk of CVD, respectively, with significant dose-response relationships identified across categories (all *P* for trend < 0.001). Similar associations were consistently confirmed across all CVD components **(Table 3**). Of note, participants with multiple CHIP faced a 2.2-fold increased risk of incident HF compared to non-carriers [HR = 2.20 (1.65-2.92), *P* < 0.001]. When restricted to CHIP carriers, higher VAF level as a continuous variable was also significantly associated with an elevated risk of CVD and CVD components (**Table S12**, all *P* < 0.05). Subsequently, we further characterized the risk profiles using the CHRS. Compared with non-carriers, participants in the high-risk category (CHRS ≥ 12.5) had a nearly two-fold increased risk of incident CVD [HR = 1.97 (1.44-2.69), *P* < 0.001], while each one-point increase in the CHRS was associated with a 14% higher risk of CVD among CHIP carriers [HR = 1.14 (1.10-1.19), *P* < 0.001]. Similar findings were consistently observed for CVD components (**Figure S3, Table S13**).

**Figure 2.**
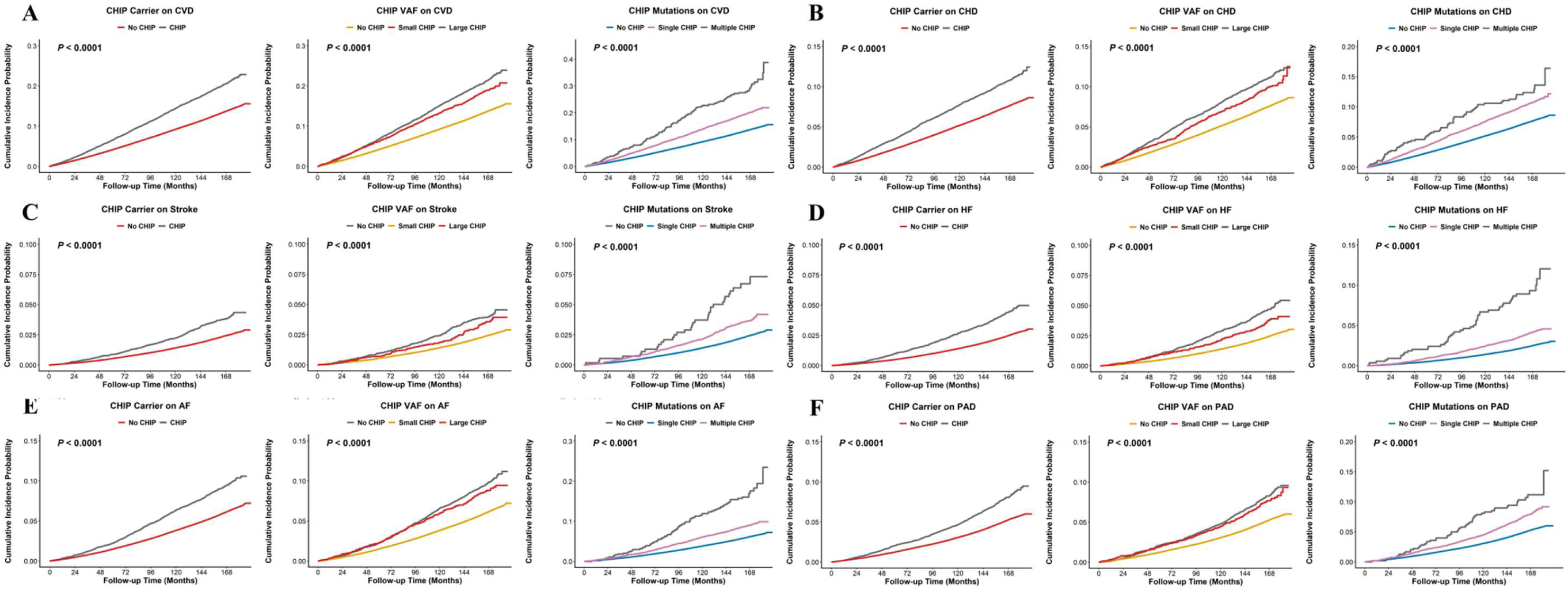
Cumulative incidence of CVD and CVD components by CHIP status. (A) Composite CVD. (B) CHD. (C) Stroke. (D) HF. (E) AF. (F) PAD. CHIP, clonal hematopoiesis of indeterminate potential; VAF, variant allele fraction; CVD, cardiovascular disease; CHD, coronary heart disease; HF, heart failure; AF, atrial fibrillation; PAD, peripheral artery disease.

**Table 3.** Association of CHIP status with the risk of CVD and specific CVD components.

| CVD and CVD components |  | No CHIP | CHIP carrier |  | CHIP VAF |  |  |  |  | CHIP mutations |  |  |  |  |
| --- | --- | --- | --- | --- | --- | --- | --- | --- | --- | --- | --- | --- | --- | --- |
|  |  |  |  |  | Small CHIP (2%≤ VAF <10%) |  | Large CHIP (VAF ≥ 10%) |  |  | Single CHIP |  | Multiple CHIP |  |  |
|  |  |  |  |  | HR (95% CI) | P value | HR (95% CI) | P value | P for trend | HR (95% CI) | P value | HR (95% CI) | P value | P for trend |
| CVD | No. cases/Total | 39176/296909 | 1976/10116 |  | 610/3394 |  | 1366/6722 |  |  | 1824/9568 |  | 152/548 |  |  |
|  | Model1 |  | 1.16 (1.11-1.22) | <0.001 | 1.09 (1.01-1.19) | 0.026 | 1.19 (1.13-1.26) | <0.001 | <0.001 | 1.14 (1.09-1.20) | <0.001 | 1.49 (1.27-1.75) | <0.001 | <0.001 |
|  | Model2 |  | 1.13 (1.08-1.18) | <0.001 | 1.07 (0.99-1.16) | 0.093 | 1.16 (1.10-1.23) | <0.001 | <0.001 | 1.11 (1.06-1.17) | <0.001 | 1.42 (1.21-1.66) | <0.001 | <0.001 |
|  | Model3 |  | 1.13 (1.08-1.18) | <0.001 | 1.08 (1.00-1.17) | 0.052 | 1.15 (1.09-1.21) | <0.001 | <0.001 | 1.11 (1.06-1.16) | <0.001 | 1.38 (1.17-1.61) | <0.001 | <0.001 |
| CHD | No. cases/Total | 21784/296909 | 1032/10116 |  | 321/3394 |  | 711/6722 |  |  | 967/9568 |  | 65/548 |  |  |
|  | Model1 |  | 1.14 (1.07-1.21) | <0.001 | 1.07 (0.96-1.20) | 0.225 | 1.17 (1.08-1.26) | <0.001 | <0.001 | 1.13 (1.06-1.21) | <0.001 | 1.19 (0.93-1.52) | 0.162 | <0.001 |
|  | Model2 |  | 1.11 (1.04-1.18) | 0.001 | 1.05 (0.94-1.17) | 0.4 | 1.14 (1.05-1.22) | <0.001 | <0.001 | 1.10 (1.04-1.18) | 0.003 | 1.15 (0.90-1.47) | 0.26 | 0.002 |
|  | Model3 |  | 1.10 (1.04-1.18) | 0.002 | 1.06 (0.95-1.18) | 0.329 | 1.13 (1.05-1.22) | 0.002 | 0.001 | 1.10 (1.03-1.18) | 0.003 | 1.12 (0.88-1.43) | 0.345 | 0.002 |
| Stroke | No. cases/Total | 6814/296909 | 365/10116 |  | 111/3394 |  | 254/6722 |  |  | 332/9568 |  | 33/548 |  |  |
|  | Model1 |  | 1.20 (1.08-1.34) | <0.001 | 1.12 (0.93-1.35) | 0.246 | 1.25 (1.10-1.41) | <0.001 | <0.001 | 1.17 (1.05-1.30) | 0.006 | 1.78 (1.27-2.51) | <0.001 | <0.001 |
|  | Model2 |  | 1.18 (1.06-1.31) | 0.003 | 1.10 (0.91-1.32) | 0.332 | 1.21 (1.07-1.38) | 0.002 | 0.002 | 1.14 (1.02-1.27) | 0.019 | 1.70 (1.20-2.39) | 0.002 | <0.001 |
|  | Model3 |  | 1.17 (1.06-1.30) | 0.003 | 1.10 (0.91-1.33) | 0.322 | 1.21 (1.07-1.37) | 0.003 | 0.002 | 1.14 (1.02-1.27) | 0.02 | 1.66 (1.18-2.34) | 0.004 | <0.001 |
| HF | No. cases/Total | 6963/296909 | 410/10116 |  | 116/3394 |  | 294/6722 |  |  | 362/9568 |  | 48/548 |  |  |
|  | Model1 |  | 1.27 (1.15-1.40) | <0.001 | 1.10 (0.92-1.32) | 0.302 | 1.35 (1.20-1.52) | <0.001 | <0.001 | 1.20 (1.08-1.33) | <0.001 | 2.42 (1.82-3.22) | <0.001 | <0.001 |
|  | Model2 |  | 1.23 (1.11-1.35) | <0.001 | 1.07 (0.89-1.29) | 0.46 | 1.30 (1.16-1.46) | <0.001 | <0.001 | 1.16 (1.04-1.28) | 0.008 | 2.27 (1.71-3.02) | <0.001 | <0.001 |
|  | Model3 |  | 1.22 (1.11-1.35) | <0.001 | 1.08 (0.90-1.30) | 0.41 | 1.29 (1.15-1.45) | <0.001 | <0.001 | 1.15 (1.04-1.28) | 0.008 | 2.20 (1.65-2.92) | <0.001 | <0.001 |
| AF | No. cases/Total | 17326/296909 | 891/10116 |  | 281/3394 |  | 610/6722 |  |  | 806/9568 |  | 85/548 |  |  |
|  | Model1 |  | 1.11 (1.04-1.19) | 0.002 | 1.08 (0.96-1.21) | 0.211 | 1.13 (1.04-1.22) | 0.004 | 0.002 | 1.07 (1.00-1.15) | 0.057 | 1.70 (1.37-2.10) | <0.001 | <0.001 |
|  | Model2 |  | 1.10 (1.02-1.17) | 0.008 | 1.06 (0.95-1.20) | 0.304 | 1.11 (1.02-1.20) | 0.011 | 0.006 | 1.06 (0.98-1.13) | 0.125 | 1.68 (1.36-2.08) | <0.001 | <0.001 |
|  | Model3 |  | 1.09 (1.02-1.17) | 0.013 | 1.07 (0.95-1.20) | 0.295 | 1.10 (1.02-1.19) | 0.019 | 0.011 | 1.05 (0.98-1.13) | 0.155 | 1.64 (1.32-2.03) | <0.001 | <0.001 |
| PAD | No. cases/Total | 14730/296909 | 759/10116 |  | 243/3394 |  | 516/6722 |  |  | 706/9568 |  | 53/548 |  |  |
|  | Model1 |  | 1.17 (1.09-1.26) | <0.001 | 1.14 (1.01-1.30) | 0.037 | 1.19 (1.09-1.29) | <0.001 | <0.001 | 1.16 (1.07-1.25) | <0.001 | 1.37 (1.05-1.80) | 0.021 | <0.001 |
|  | Model2 |  | 1.13 (1.05-1.22) | <0.001 | 1.12 (0.99-1.27) | 0.075 | 1.14 (1.04-1.24) | 0.004 | 0.001 | 1.12 (1.04-1.21) | 0.003 | 1.29 (0.98-1.69) | 0.065 | <0.001 |
|  | Model3 |  | 1.13 (1.05-1.22) | 0.001 | 1.13 (1.00-1.28) | 0.06 | 1.13 (1.04-1.24) | 0.006 | 0.002 | 1.12 (1.04-1.21) | 0.003 | 1.27 (0.97-1.66) | 0.083 | <0.001 |

*Abbreviations:* CHIP, clonal hematopoiesis of indeterminate potential; CVD, cardiovascular disease; CHD, coronary heart disease; HF, heart failure; AF, atrial fibrillation; PAD, peripheral artery disease; HR, hazard ratio; CI, confidence interval; VAF, variant allele fraction.
All hazard ratios are presented in comparison to the participants with No CHIP .
Model 1 was adjusted for age, sex, and ethnicity.
Model 2 was further adjusted for BMI, education, Townsend deprivation index, smoking status, alcohol consumption, sleep duration, physical activity, diet quality, and assessment center.
Model 3 was further adjusted for hypertension, diabetes, and use of antihypertensive, antidiabetic, and lipid-lowering medications.

Although subgroup analyses (**Figure S4, Table S14**) demonstrated that the association between CHIP status and CVD risk was generally consistent across most strata, a significant interaction was observed for age (*P* for interaction < 0.05). The impact of CHIP appeared more pronounced in younger participants compared to older participants. Additionally, CHIP showed a tendency toward a stronger adverse effect among individuals with a history of alcohol consumption (*P* for interaction < 0.05). Sensitivity analyses (**Table S15**) confirmed the robustness of these findings, as the significant associations persisted after excluding cases diagnosed within the first 2 or 5 years of follow-up, excluding early deaths, excluding participants with urinary albumin levels below the detection limit, or further adjusting for the PREVENT 10-year CVD risk score.

### 3.4 Gene-specific CHIP and CVD risk

Gene-specific analyses revealed distinct risk profiles across different driver mutations (**Figure S5, Table S16**). Regarding composite CVD events, carriers of *PRPF8* CHIP faced the highest CVD risk [HR 2.41 (1.37–4.25), *P* = 0.002], followed by mutations in *JAK2* [HR 2.09 (1.45–3.03), *P* < 0.001], *SF3B1* [HR 1.76 (1.21–2.57), *P* = 0.003], *SRSF2* [HR 1.63 (1.24–2.12), *P* < 0.001], *PPM1D* [HR 1.27 (1.00–1.61), *P* = 0.046], and *TET2* [HR 1.20 (1.08–1.34), *P* < 0.001]. The associations varied substantially across CVD components. The most prevalent mutation, such as *TET2* CHIP, was consistently associated with elevated risks of CHD, stroke, and AF. In contrast, *ASXL1* CHIP showed stronger associations with HF and PAD. However, despite being the most frequently mutated gene, *DNMT3A* CHIP was not significantly associated with most CVD outcomes after full adjustment. Notably, *PRPF8* CHIP conferred the highest risk for CHD and AF, while *SF3B1* CHIP contributed the highest risk for stroke, HF, and PAD. These associations were largely preserved and more pronounced among carriers of large CHIP; for example, the risk of HF associated with any *TP53* CHIP was 2.14-fold, reaching 2.70-fold for those with large *TP53* CHIP.

### 3.5 Joint association of CHIP status and CKM syndrome with CVD risk

Compared with participants at CKM stages 0–1 without CHIP at baseline, the risk of incident CVD demonstrated a stepwise increasing trend with advancing CKM stages, an effect that was further amplified by the presence of CHIP (**Figure 3, Table S17**). A significant dose-response relationship was observed across strata (*P* for trend < 0.001), with the highest CVD risk reached among those at CKM stage 3 with CHIP [HR = 1.63 (1.50-1.78), *P* < 0.001]. Notably, even at early CKM stages 0–1, participants with CHIP exhibited a numerically higher risk of CVD [HR = 1.36 (1.10-1.69), *P* = 0.005] than those at CKM stage 2 without CHIP [HR = 1.25 (1.18-1.33), *P* < 0.001]. When further stratified by CHIP VAF and mutations, this dose-response gradient became even more pronounced (all *P* for trend < 0.001). The highest CVD risks were observed in CKM Stage 3 participants with large CHIP [HR 1.64 (1.49-1.80), *P* < 0.001] or with multiple CHIP [HR 1.94 (1.58-2.37), *P* < 0.001].

**Figure 3.**
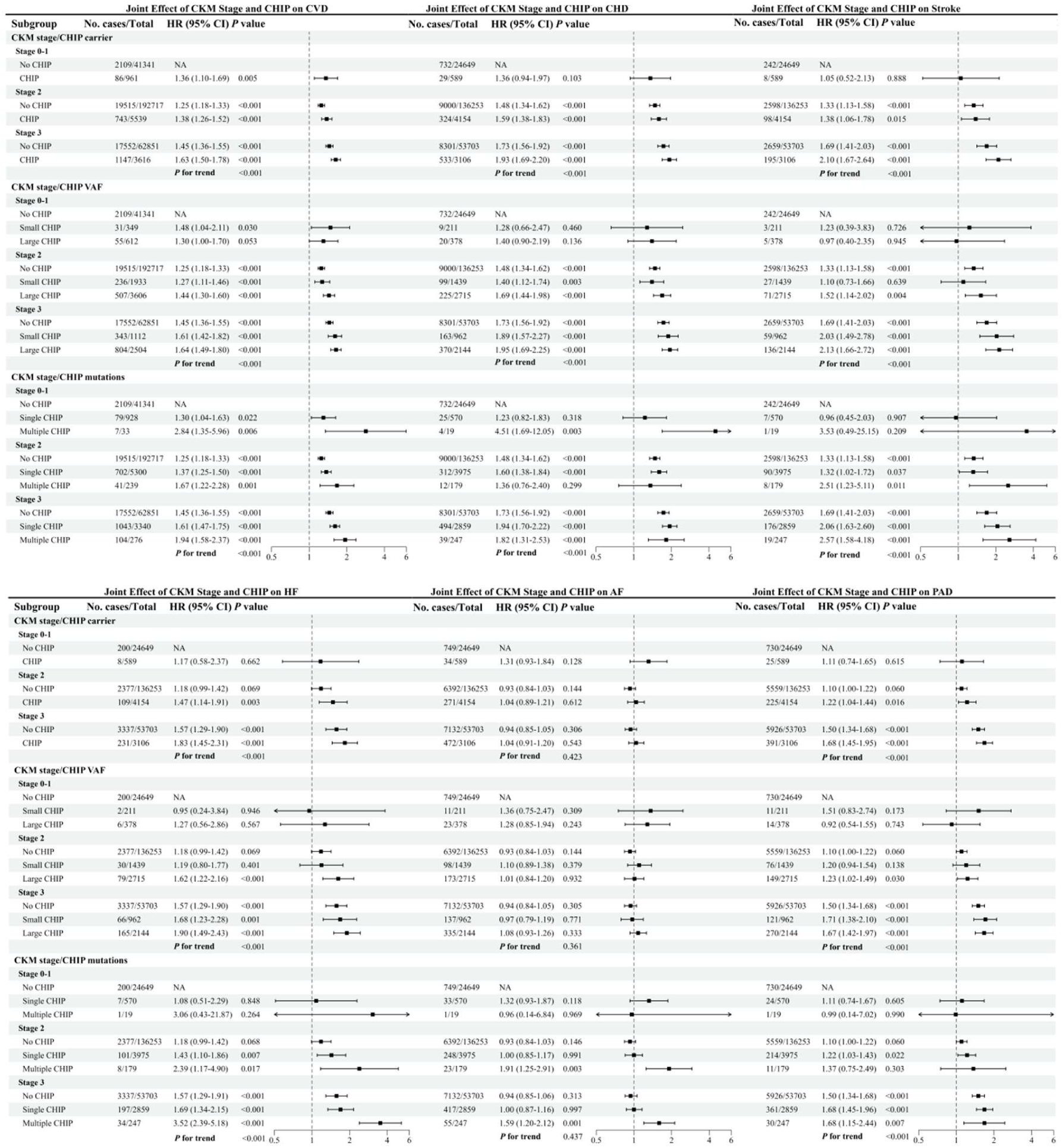
Joint associations of CHIP status and CKM syndrome stage with the risks of incident CVD and CVD components. CKM, cardiovascular-kidney-metabolic; CHIP, clonal hematopoiesis of indeterminate potential; CVD, cardiovascular disease; CHD, coronary heart disease; HF, heart failure; AF, atrial fibrillation; PAD, peripheral artery disease; HR, hazard ratio; CI, confidence interval.

These patterns remained consistent across most CVD components. The additive effect of multiple CHIP was particularly striking for HF, with a more than threefold increased risk [HR 3.17 (2.21-4.56), *P* < 0.001] observed in CKM Stage 3. Furthermore, among participants at CKM stages 0–1, the presence of multiple CHIP was associated with a sharp, discontinuous increase in the risk for composite CVD events [HR = 2.84 (1.35-5.96), *P* = 0.006], CHD [HR = 3.36 (1.26-8.98), *P* = 0.016], and HF [HR = 4.49 (1.12-18.05), *P* = 0.034], although these estimates were based on limited numbers.

### 3.6 Joint association of CHIP status and genetic risk

Based on the training set, a GWAS was conducted for CKM syndrome stage (0–3), with summary statistics provided in **Table S18**. The Manhattan plot identified genome-wide significant loci associated with CKM progression (**Figure S6A**), while the Q-Q plot indicated minimal genomic inflation (**Figure S6B**, λGC = 1.006). The constructed CKM-PRS followed a normal distribution in both sets (**Figure S6C**). The proportion of participants in more advanced CKM stages increased stepwise across ascending PRS deciles (**Figure S6D**), with standardized PRS values rising progressively from stage 0 to stage 3 (**Figure S6E**). Similarly, PRS for CVD components were constructed using external GWAS summary data (**Figure S7**).

The joint effects of genetic susceptibility and CHIP status on CKM progression and CVD risk revealed significant joint associations (all *P* for trend < 0.001). As shown in **Table S19 and Figure S8**, compared to the reference group (low genetic risk and no CHIP), participants with both high genetic risk and CHIP carrier status faced a 38% higher risk of harboring a more advanced CKM stage [OR 1.38 (1.25-1.52), *P* < 0.001]. This association was even more pronounced for those with high genetic risk combined with large CHIP [OR 1.41 (1.25-1.59), *P* < 0.001] or multiple CHIP [OR 1.87 (1.24-2.82), *P* = 0.003].

This additive pattern was consistently replicated for CVD components (**Table S20,** all *P* for trend < 0.001). The combination of high genetic risk and CHIP significantly increased the risk of CHD [HR 1.32 (1.21-1.44), *P* < 0.001], stroke [HR 1.39 (1.21-1.60), *P* < 0.001], HF [HR 1.31 (1.14-1.51), *P* < 0.001], AF [HR 1.40 (1.28-1.54), *P* < 0.001], and PAD [HR 1.19 (1.07-1.31), *P* = 0.001]. Similarly, high genetic risk combined with large CHIP or multiple CHIP consistently conferred the greatest risk across most endpoints.

### 3.7 Incremental predictive value of CHIP beyond the PREVENT score

As shown in **Figure 4**, CHIP status provided additional stratification of CVD risk across tertiles, quintiles, and deciles of the PREVENT score. Consistently, all extended models incorporating CHIP-related variables showed statistically significant improvements in both the NRI and IDI (all *P* < 0.05) (**Table S21**).

**Figure 4.**
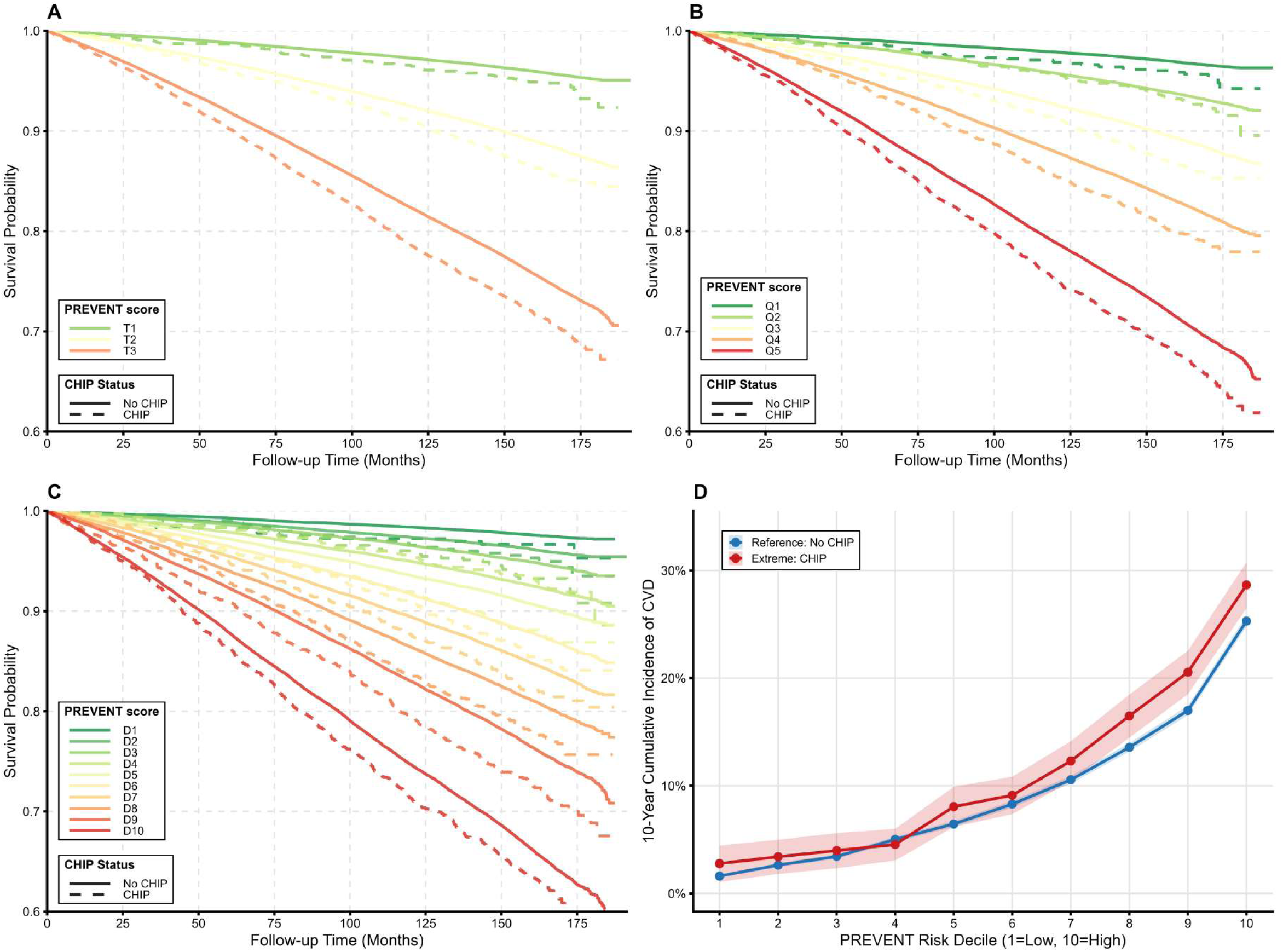
Risk stratification for incident CVD by PREVENT scores and any CHIP status. (A) Tertiles. (B) Quintiles. (C) Deciles. (D) 10-year cumulative incidence of CVD across PREVENT deciles stratified by any CHIP. CHIP, clonal hematopoiesis of indeterminate potential; CVD, cardiovascular disease.

## 4. Discussion

In this prospective cohort study, we systematically investigated the association of CHIP with incident CVD risk within the CKM syndrome framework. The principal findings are as follows: (1) CHIP carriers (particularly those with large CHIP or multiple CHIP) were more likely to present with advanced CKM stages at baseline. (2) CHIP significantly increased the risk of incident CVD, with distinct risk gradients according to clonal burden and driver gene subtype. (3) CHIP and CKM stages jointly shaped CVD risk. (4) The coexistence of CHIP and high genetic risk further increased susceptibility to CVD. (5) Incorporating CHIP-related variables significantly improved the risk discrimination of the current PREVENT score, supporting the utility of CHIP for refining cardiovascular risk stratification in the CKM framework.

CHIP has emerged as a novel cardiovascular risk factor, supported by a wealth of clinical and experimental evidence over recent years. Population-based studies consistently demonstrate that CHIP carriers face significantly elevated risks of major CVD events and those harboring large or multiple CHIP exhibit even greater hazards, following a clear dose-response relationship ^[9,25]^. Our study further corroborates this association within a CKM stage 0–3 population free of baseline CVD, observing that CHIP status was associated with a 13% increase in incident composite CVD risk, with large and multiple CHIP conferring approximately 15% and 38% excess risks, respectively. Mechanistically, CHIP accelerates atherosclerotic progression and myocardial injury by driving chronic inflammation and immune dysregulation ^[8,26,27].^ Notably, distinct driver genes exhibit significant heterogeneity in their impact on CVD components. *TET2* mutations, by hyperactivating the NLRP3–IL-1β/IL-6 inflammasome axis ^[26,28]^, and ASXL1 mutations, potentially acting through IRAK1/TAK1–NF-κB or AIM2 pathways ^[29,30]^, both potentiate the pro-inflammatory phenotype of monocytes/macrophages, culminating in endothelial injury, increased plaque burden and instability. Animal models further confirm that inhibiting IL-1β/IL-6 signaling partially attenuates CHIP-mediated CVD risk, underscoring the pivotal role of inflammatory pathways ^[31]^. Moreover, *JAK2* mutations promote coronary and venous thrombosis as well as microvascular dysfunction ^[8,9]^, while DNA damage repair genes (e.g., *TP53, PPM1D*) and spliceosome genes (e.g., *SF3B1, SRSF2, PRPF8*) are associated with systemic atherosclerosis burden across multiple vascular beds and adverse cardiovascular outcomes ^[32]^. Consistent with prior studies, we found that *TET2* was persistently associated with elevated risks of CHD, stroke, and AF, whereas *ASXL1* exhibits a stronger predilection for HF and PAD. In contrast, despite being the most prevalent mutation, *DNMT3A* was not significantly associated with most CVD outcomes after full adjustment for traditional risk factors, aligning with findings from large cohorts suggesting that *DNMT3A* confers a milder vasculotoxic potential compared to *TET2* or *JAK2* ^[25,33]^. Furthermore, we identified that spliceosome genes such as *SF3B1*, *SRSF2*, and *PRPF8* conferred some of the highest relative risks for specific CVD components, resonating with observations from an ASCVD cohort and suggesting that aberrant RNA splicing may represent an important novel pathway driving CHIP-mediated multi-vascular bed pathology and myocardial remodeling ^[25]^.

Our findings position CHIP not merely as a biomarker of biological aging but as a crucial driver accelerating the progression of CKM syndrome from its earliest stages. CHIP, particularly mutations in *TET2* and *ASXL1*, has been associated with an elevated risk of incident type 2 diabetes. Experimental evidence demonstrates that *TET2* clonal expansion in diet-induced obese mice significantly exacerbates obesity-related insulin resistance and hyperglycemia, effects likely mediated by potentiated IL-1β expression in white adipose tissue ^[34]^. Similarly, obesity appears more prevalent among CHIP carriers ^[35]^, with prevalence increasing progressively alongside waist-to-hip ratio ^[36]^, further suggesting a link between CHIP and adverse metabolic states. Moreover, CHIP—especially non-*DNMT3A* mutations (e.g., *TET2*, *ASXL1*)—is significantly associated with CKD, acute kidney injury (AKI), and a more rapid decline in eGFR ^[37,38]^. In CKD mouse models, *TET2* CHIP leads to more severe renal tubular injury, interstitial inflammation, and fibrosis ^[38]^. Concurrently, conditions such as obesity, chronic inflammation, and CKD can promote the expansion of CHIP clones via signaling pathways like IFN-γ and TNFα, creating a vicious cycle that accelerates CKM progression ^[39]^. Crucially, our data show that individuals in early CKM stages (0–1) carrying CHIP face a significantly higher CVD risk than those in a more advanced stage (stage 2) without CHIP. This phenomenon of possible risk crossover suggests that current metabolic-centric CKM frameworks, including the PREVENT equations, may systematically underestimate the residual inflammatory risk primarily driven by CHIP. Incorporating CHIP into prediction models significantly improved risk discrimination, correctly reclassifying ∼4% of the population despite a CHIP prevalence of only 3.3%—a notable gain, especially compared with adding lipoprotein(a) to PREVENT, which yielded an NRI of only 0.006 ^[40]^.

Our study has several limitations. First, UK Biobank participants are predominantly of European ancestry and exhibit a healthy volunteer bias. Furthermore, the exclusion of participants with missing variables may introduce selection bias, potentially limiting the generalizability and representativeness of our findings. Second, CHIP status was assessed only once at baseline, precluding the assessment of dynamic evolution of clones over time. Third, compared with deep targeted sequencing, the limited sequencing depth of WES may have resulted in an underestimation of CHIP prevalence and the specific impact of small clones. Fourth, although our sample size was substantial, validation in independent cohorts would further strengthen the robustness of our conclusions. Fifth, despite comprehensive adjustments and sensitivity analyses excluding early-onset cases or early deaths, we cannot entirely rule out the possibility of reverse causality or residual confounding.

## 5. Conclusions

Our study confirms CHIP as a risk factor associated with more advanced CKM stages and amplifies CVD risk. Integrating CHIP into existing CVD prevention strategies could significantly optimize risk stratification.

## Data Availability

We performed analyses based on UKB and publicly available GWAS summary statistics.

## List of abbreviations

AF: Atrial fibrillation
AHA: American Heart Association
BMI: Body Mass Index
CHD: Coronary heart disease
CHIP: Clonal hematopoiesis of indeterminate potential
CHRS: Clonal Hematopoiesis Risk Score
CI: Confidence intervals
CKD: Chronic kidney disease
CKM: Cardiovascular-kidney-metabolic
CVD: Cardiovascular disease
eGFR: Estimated glomerular filtration rate
GWAS: Genome-wide association studies
HF: Heart failure
HR: Hazard ratios
ICD-10: International Classification of Diseases, Tenth Revision
IDI: Integrated Discrimination Improvement
IL: Interleukin
KDIGO: Kidney Disease: Improving Global Outcomes
LD: Linkage disequilibrium
MAC: Minor allele count
MAF: Minor allele frequency
MREC: Multi-centre Research Ethics Committee
NRI: Net Reclassification Improvement
OR: Odds ratios
PAD: Peripheral artery disease
PREVENT: Predicting Risk of CVD EVENTs
PRS: Polygenic risk scores
QC: Quality control
Q-Q: Quantile-Quantile
TDI: Townsend Deprivation Index
WES: Whole-exome sequencing
WGS: Whole-genome sequencing

## Supplementary Information

**Additional file 1. Supplementary Figures** (**Figure S1**. Flowchart of this study. **Figure S2**. Age-stratified distribution of CHIP status across CKM syndrome stages. **Figure S3**. Association of CHRS with risks of CVD and CVD components. **Figure S4**. Subgroup analysis of the association between CHIP status and CVD risk. **Figure S5**. Association of gene-specific CHIP carrier with the risk of CVD and CVD components. **Figure S6**. GWAS results of CKM syndrome and characteristic of CKM – PRS. **Figure S7**. Characteristic of CVD components – PRS. **Figure S8**. Joint associations of genetic risk and CHIP status with more advanced baseline CKM stages.)

**Additional file 2. Supplementary Tables** (**Table S1.** Summary of Field IDs and missing information for variables defining CKM syndrome in UK Biobank. **Table S2.** Detailed information of all GWAS datasets used in this study. **Table S3**. Definition of stages of CKM syndrome in the UK Biobank. **Table S4.** AHA PREVENT equations for 10-year CVD risk estimation. **Table S5**. KDIGO risk stratification criteria for CKD. **Table S6**. CHIP related genes and mutations used to identify CHIP status. **Table S7**. Genetic and clinical characteristics used in the clonal hematopoiesis risk score (CHRS) with corresponding score weights. **Table S8**. Construction of healthy diet score from UK Biobank baseline questionnaire. **Table S9**. Codes used to identify drug classification. **Table S10**. Codes used to identify baseline comorbidities and prevalent cardiovascular outcomes. **Table S11**. Baseline characteristics of participants according to CHIP status. **Table S12**. Association of CHIP VAF as a continuous variable with the risk of CVD and CVD components among CHIP carriers. **Table S13**. Association of CHRS with the risk of CVD and CVD components. **Table S14**. Subgroup analysis of the association of CHIP status with the risk of CVD. **Table S15**. Sensitivity analyses of the association between CHIP status and CVD. **Table S16**. Association of gene-specific CHIP carrier with the risk of CVD and CVD components. **Table S17.** Joint associations of CHIP status and CKM syndrome stage with the risks of incident CVD and CVD components. **Table S18.** GWAS summary statistics for CKM syndrome stage (0–3) in the training set. **Table S19**. Joint associations of genetic risk and CHIP status with the risk of more advanced CKM syndrome stages. **Table S20**. Joint associations of genetic risk and CHIP status with the risks of incident CVD components. **Table S21**. Incremental predictive value of adding CHIP related variable to the PREVENT risk score for 10-year incident CVD.)

## Acknowledgements

Not applicable.

## Ethics approval and consent to participate

The UKB study received ethical approval from the North West Multi-Centre Research Ethics Committee (21/NW/0157). In accordance with the principles outlined in the Helsinki Declaration, written informed consent was obtained from all participants prior to their inclusion in the study.

## Consent for publication

Not applicable.

## Availability of data and materials

We performed analyses based on UKB and publicly available GWAS summary statistics.

## Authors’ contributions

Jian Lu: Conceptualization, Writing - review & editing, Investigation, Data curation, Funding acquisition, Validation. Shuaigang Sun: Writing - original draft, Methodology, Formal analysis, Visualization, Software. Zekai Deng: Methodology. Shunwei Wang: Methodology. Chenping Wei: Methodology. Shimin Jiang: Funding acquisition, Project administration. Wenge Li: Funding acquisition, Supervision. Jian Lu and Shuaigang Sun contributed equally to this work. All authors read and approved the final manuscript.

